# Clinician Perspectives on Automated Facial Analysis for Pain Assessment: A Cross-Sectional Survey of Physicians, Advanced Practice Providers, and Nurses

**DOI:** 10.64898/2026.09.21.26363597

**Authors:** Colleen S. Mullins, Cori Espelein, Matthew J. Gurka, Pavel Chernyavsky

**Author notes:** **Corresponding author:** Colleen Mullins, School of Medicine, University of Virginia, PO Box 800793, Charlottesville, VA 22903.

## Abstract

Pain assessment remains challenging in clinical practice. Automated pain assessment using artificial intelligence (AI) and facial expression analysis has emerged as a potential approach to support more objective and continuous pain monitoring, particularly for patients who are unable to communicate effectively. However, successful implementation depends on clinician acceptance. We conducted a cross-sectional survey of physicians, advanced practice providers (APPs), and nurses at a tertiary academic medical center to evaluate current pain assessment practices and perceptions of a facial-analysis-based pain assessment system. Eighty-seven eligible clinicians participated. For current assessment practices, clinicians commonly relied on observational indicators of pain, including vital signs (88.5%), physical examination findings (85.1%), and facial expressions (79.3%). Attitudes toward automated pain assessment were generally neutral to positive. Half of respondents agreed that such a system would be useful for monitoring patients over time (50.6%), and many perceived value for nonverbal patients (86.5%), patients with dementia or cognitive impairment (78.4%), and patients with language barriers (62.2%). Most clinicians reported requiring high levels of accuracy before routine use, with nearly 60% requiring at least 90% accuracy. Key concerns included alert fatigue and unequal performance across patient populations. These findings suggest clinicians view automated facial-analysis systems as potentially valuable adjuncts for pain monitoring, particularly in communication-impaired populations, while emphasizing the importance of accuracy, fairness, explainability, and workflow integration during system development and implementation.

## Introduction

Pain is the most common reason patients seek medical attention.^1^ Accurate pain assessment is essential for guiding appropriate treatment; yet, current methods remain limited in their ability to translate the patient’s subjective experience into an objective clinical measure.^2^ Commonly used tools, such as the Numeric Rating Scale (NRS) and Visual Analog Scale (VAS), require patients to reduce a multidimensional experience to a single point on a scale, and have not significantly improved the timeliness or quality of pain management.^2,3^ These scales are further limited in populations unable to self-report, including patients with cognitive impairment or dementia, nonverbal patients, and young pediatric populations.^4,5^

Pain reassessment, when analgesic dosage can be adjusted or alternative strategies initiated, is often performed less frequently than recommended. A study across five intensive care units (ICUs) found a median of only three documented pain assessments per 24-hour period, well below the recommended eight to twelve assessments.^6^ Other studies have noted poor compliance with reassessment standards following analgesic administration,^7^ in both primary care^8^ and acute care settings.^9^ Continuous monitoring of pain by clinicians is not feasible due to workload and clinical workflow constraints. Observer-based pain evaluation is also susceptible to biases related to clinicians’ personal beliefs, the patient-provider relationship, and even physical attractiveness.^10–12^ Automatic pain recognition and scoring systems may help address these gaps by enabling more objective and continuous assessment.^13^

Facial expressions provide an intuitive social signal of pain in humans.^14,15^ Most research on facial expression analysis is grounded in the Facial Action Coding System (FACS), which decomposes facial expressions into 46 elementary muscle groups or “action units” (AUs).^16^ A systematic review of studies using FACS to analyze facial activity during pain identified a consistent subset of pain-related AUs across experimental and clinical pain, different pain modalities, and diverse patient populations, including individuals with mild cognitive impairment.^17,18^ Emerging artificial intelligence (AI) models can track these facial actions automatically, with recent studies reporting promising diagnostic accuracy for pain detection from facial images.^19–21^

Successful integration of automated pain recognition into clinical practice depends critically on clinician acceptance and perceived utility.^22^ A survey of anesthetists and intensive care nurses found that 50% would use automated pain recognition technology.^23^ More broadly, research on AI adoption in healthcare has demonstrated that clinicians generally recognize the potential benefits of AI, but barriers include concerns regarding trust, accuracy, interpretability, and workflow integration.^24–26^ These findings underscore the need to assess clinician perspectives across diverse clinical settings to guide effective application of automated pain recognition.

We conducted a survey study to understand clinicians’ perspectives on challenges in assessing pain, perceived barriers to adoption of an automatic pain assessment system, and willingness to use facial analysis technology for pain assessment. Addressing these questions is essential, as interdisciplinary collaboration and clinician engagement throughout the development process is crucial for successful AI integration in clinical practice.

## Methods

### Study Design and Setting

We conducted a cross-sectional survey of physicians, advanced practice providers (APPs), and nurses at an academic tertiary care center in the South Atlantic US state. The survey is part of a larger parallel mixed-methods study that also includes semi-structured qualitative interviews with emergency department clinicians, reported separately. The broader study was designed to collect cross-sectional input from healthcare providers to inform the development of clinician-facing AI tools for pain assessment. The Checklist for Reporting Results of Internet E-Surveys (CHERRIES)^27^ (See Supplementary Material 1) guidelines were followed to ensure comprehensive reporting of the research processes and outcomes.

### Survey Development and Administration

A survey instrument was developed to collect: 1) clinician background characteristics, including clinical role, department, years in clinical role, age, and gender; 2) current pain assessment practices; 3) challenges and limitations in assessing patient pain; 4) perceived benefits and concerns regarding a facial analysis-based pain assessment tool; and 5) willingness to use such technology in clinical practice. The survey comprised 6 multiple choice, 4 select all that apply, and 24 Likert scale questions (5 levels), as well as two opportunities for free-text comments regarding benefits, trust, and concerns about the technology (See Supplementary Material 2 for full survey). These items were included to capture concerns not represented in the closed-ended items and to contextualize quantitative responses. All questions were non-mandatory and could be skipped. Question development was guided by a review of literature on current pain assessment practices and physician trust in AI.^23,28,29^ The survey was administered on the Qualtrics platform (Qualtrics, Provo, UT). Because an anonymous link was used, the survey could not prevent multiple responses from a singular participant, although the risk was considered low due to lack of participation incentives.

### Study Participants and Recruitment

Eligible participants for the survey included physicians, advanced practice providers, and registered nurses working in the following clinical departments and units: Emergency Department, Intensive Care Unit (ICU), Surgical Services (including Orthopedic Surgery and Trauma Surgery), Anesthesiology, Post-Anesthesia Care Unit (PACU), Oncology, Palliative Care, Family Medicine, Pediatrics, and Inpatient Medicine. Participants were recruited using convenience sampling. A survey link was distributed by nurse managers and department chairs via institutional email to eligible clinicians across the specified departments beginning June 16, 2026. The survey remained open for responses through August 3, 2026. Participation was voluntary and anonymous. Because the exact number of individuals who received the survey link could not be determined, a precise response rate could not be calculated. No formal a priori power calculation was conducted, consistent with the exploratory and descriptive nature of the study. The study aimed to collect as many responses as possible during the study period.

### Data Analysis

Survey data were downloaded from Qualtrics into R (Vienna, Austria). Descriptive statistics were calculated for all multiple choice, select all that apply, and Likert scale questions. Exploratory subgroup analyses evaluated whether response patterns differed according to clinician characteristics, including clinical role, years in clinical role (≤5 years vs. >5 years) and gender. Clinical role was categorized as physician (attending and resident physicians combined), advanced practice provider, or registered nurse. Gender-based analyses were limited to respondents identifying as men or women; due to small sample size (n=1), the non-binary category was excluded. Survey items measured on 5-point Likert scales were collapsed into three categories (disagree/strongly disagree, neutral, and agree/strongly agree) to reduce sparse cell counts. Associations between subgroup characteristics and survey responses were evaluated using two-tailed Fisher’s exact tests. Because these analyses were exploratory, no p-value adjustment for multiple comparisons was performed. Item-level analyses were conducted using available-case data, with the number of respondents contributing to each analysis reported in the corresponding results. A p-value <0.05 was considered statistically significant.

Item-level missing data varied across items and are reflected in the reported denominators alongside each result. To assess the overall pattern of missingness, the Little missing completely at random (MCAR) test was applied to survey variables included in the analytic dataset after exclusion of free-text responses and selected role-dependent or multiple-response items.^30^

Free-text survey responses were analyzed using content analysis to develop a summary of key results.^31^ All answers were extracted from Qualtrics and imported into ATLAS.ti (version 26). The text was coded independently by two members of the research team. Codes and categories were discussed between researchers; where discrepancies arose, researchers referred to the raw data and reached consensus.^31^

Within the results section, quantitative findings are presented first, followed by representative quotes from qualitative analysis that expand upon the quantitative data. The free-text responses provide additional context for the quantitative findings but should be interpreted as exploratory, as only a subset of participants provided written comments.

### Ethical Considerations

This study was approved by the University of Virginia Institutional Review Board for Social and Behavioral Sciences (IRB-SBS) under Protocol Number 8544. Informed consent was obtained from all participants prior to beginning the survey. Survey responses were collected anonymously. Respondents did not receive compensation.

## Results

### Survey Participant Characteristics and Data Completeness

A total of 90 survey responses were collected. Two responses were excluded because the respondents’ clinical roles did not meet the inclusion criteria; one response was excluded because the respondent’s department did not meet the inclusion criteria. The remaining 87 participants included 23 attending physicians, 6 resident physicians, 20 APPs, and 38 nurses. 23 respondents provided a total of 29 free-text responses. Demographic and clinical characteristics are reported in Table 1.

**Table 1.**
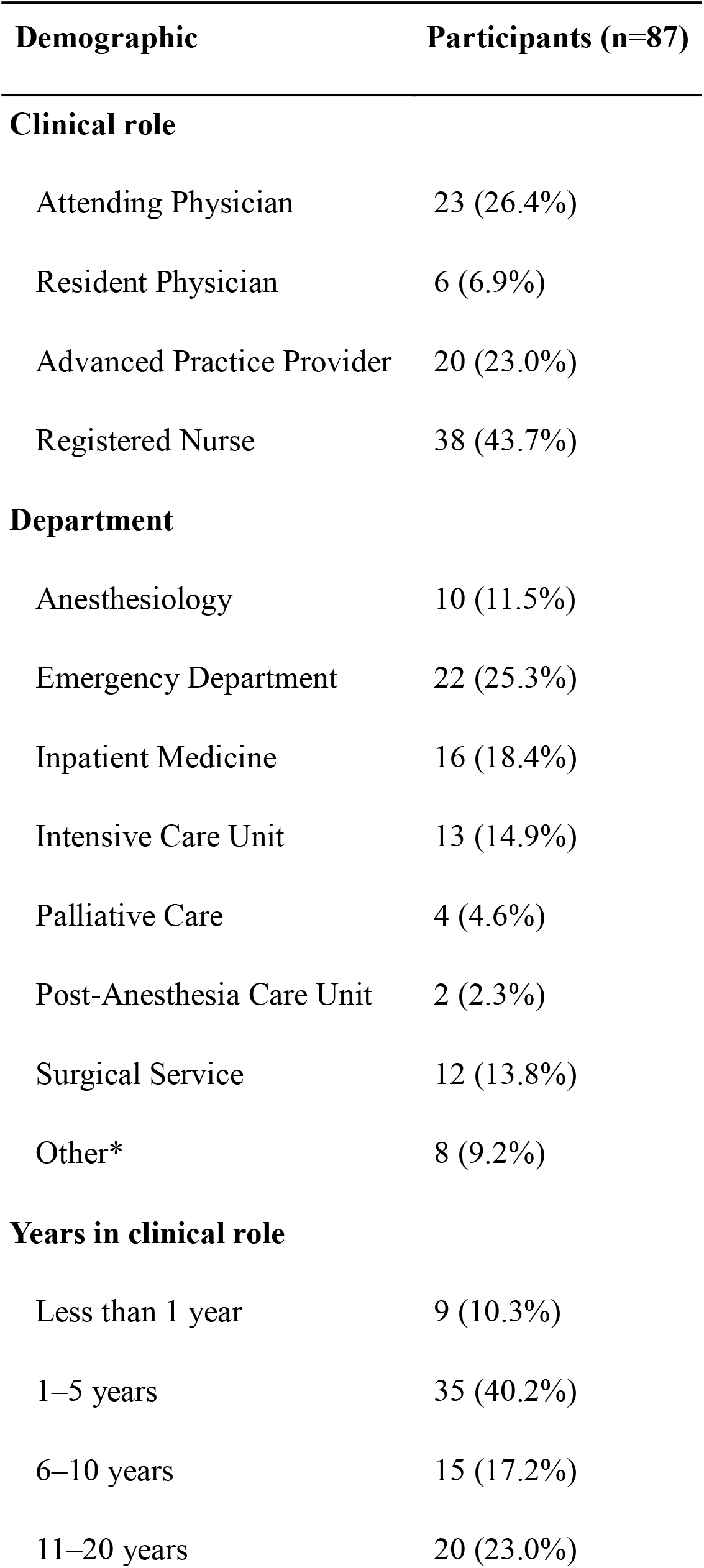
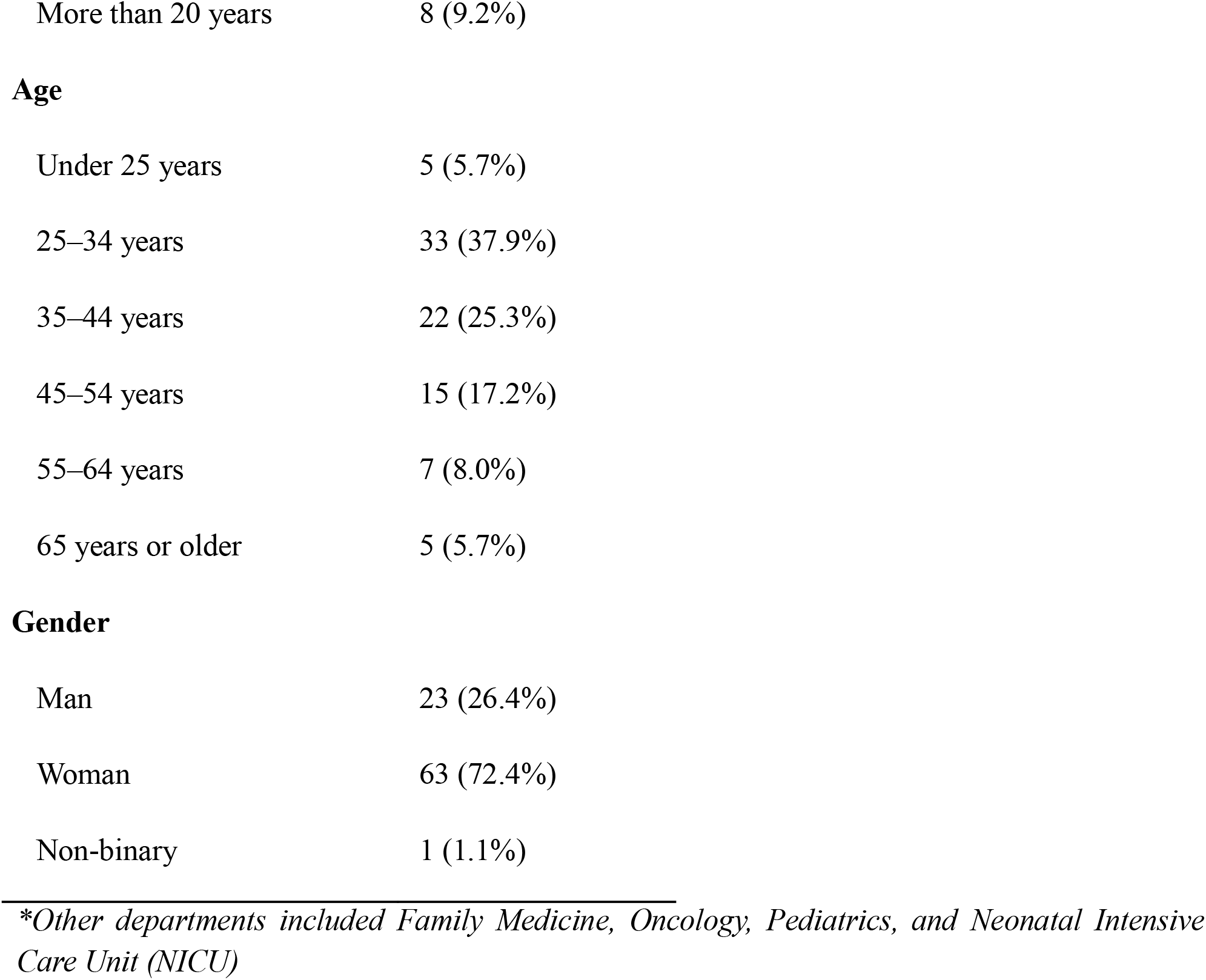
Participant demographics.

| **Demographic** | **Participants (n=87)** |
| --- | --- |
| **Clinical role** |  |
| Attending Physician | 23 (26.4%) |
| Resident Physician | 6 (6.9%) |
| Advanced Practice Provider | 20 (23.0%) |
| Registered Nurse | 38 (43.7%) |
| **Department** |  |
| Anesthesiology | 10 (11.5%) |
| Emergency Department | 22 (25.3%) |
| Inpatient Medicine | 16 (18.4%) |
| Intensive Care Unit | 13 (14.9%) |
| Palliative Care | 4 (4.6%) |
| Post-Anesthesia Care Unit | 2 (2.3%) |
| Surgical Service | 12 (13.8%) |
| Other* | 8 (9.2%) |
| **Years in clinical role** |  |
| Less than 1 year | 9 (10.3%) |
| 1–5 years | 35 (40.2%) |
| 6–10 years | 15 (17.2%) |
| 11–20 years | 20 (23.0%) |
| More than 20 years | 8 (9.2%) |
| **Age** |  |
| Under 25 years | 5 (5.7%) |
| 25–34 years | 33 (37.9%) |
| 35–44 years | 22 (25.3%) |
| 45–54 years | 15 (17.2%) |
| 55–64 years | 7 (8.0%) |
| 65 years or older | 5 (5.7%) |
| **Gender** |  |
| Man | 23 (26.4%) |
| Woman | 63 (72.4%) |
| Non-binary | 1 (1.1%) |

*\*Other departments included Family Medicine, Oncology, Pediatrics, and Neonatal Intensive Care Unit (NICU)*

The level of missing data was low overall (173/2871, 6.0% of cells). Little’s test indicated that data were not missing “completely at random” (X^2^(241) = 302.0, p = 0.004). Because the overall level of missingness was low, analyses were conducted using available-case responses for each survey item. Full survey results are reported in Supplementary Material 3, with all Fisher’s test results in Supplementary Material 4.

### Current pain assessment practices

Respondents reported using a wide range of factors to assess pain (Figure 1). Providers most frequently used vital signs (77/87, 88.5%), physical examination findings (74/87, 85.1%), facial expressions (69/87, 79.3%), and the patient’s diagnosis/injury type (55/87, 63.2%) as observational sources of information. For patient-reported sources, clinicians most often used the patient-reported NRS score (67/87, 77.0%) and verbal description of pain (67/87, 77.0%).

**Fig. 1.**
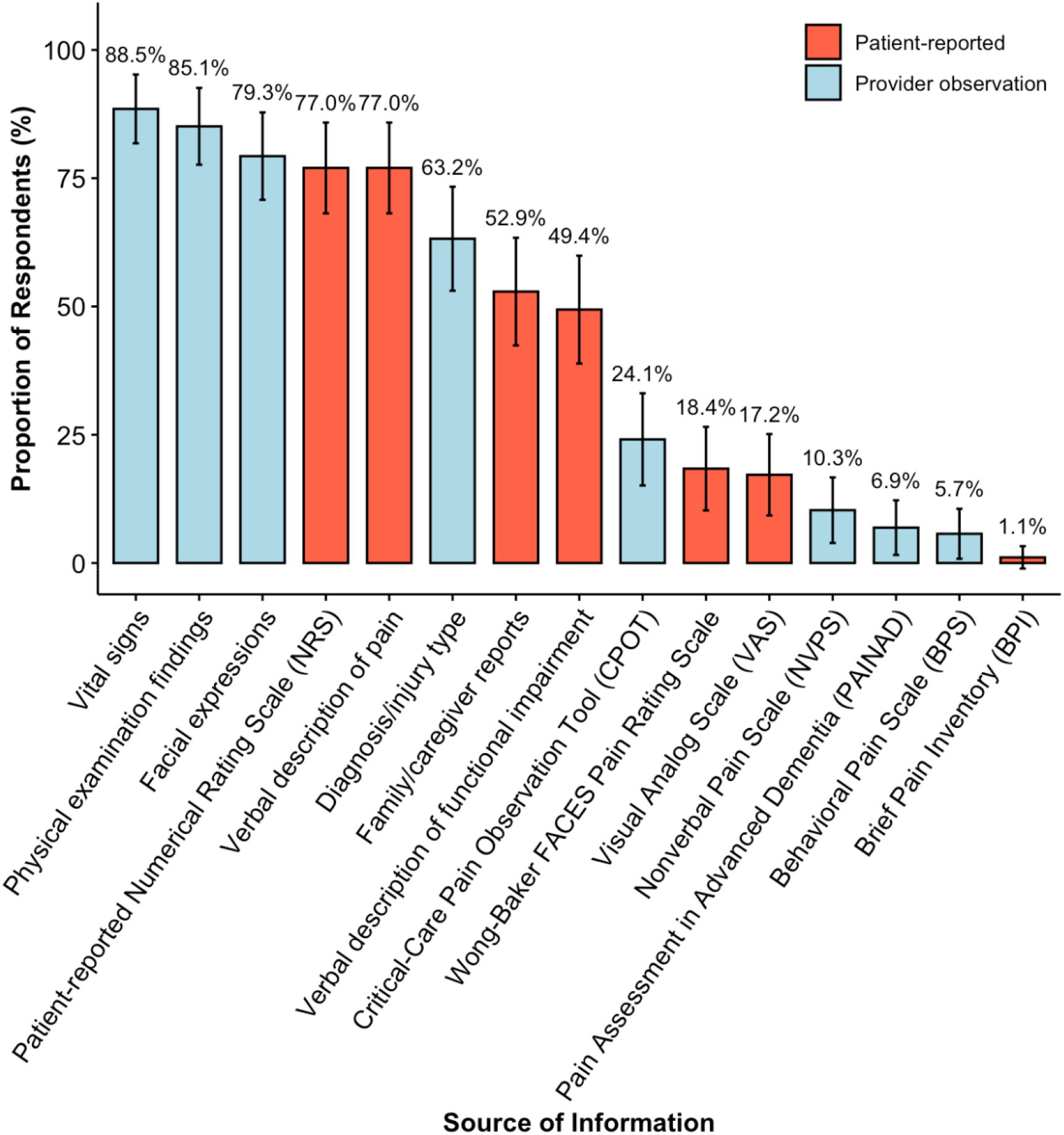
Current pain assessment tools reported by clinicians in an academic tertiary car center (N=87). Note: “Other” category excluded; participants selected all that apply. Vertical whiskers represent 95% Confidence Intervals. Figure created in R

### Perceptions on facial-analysis technology for pain assessment

Regarding the perceived clinical utility of a potential automatic pain recognition and scoring system, responses were neutral-to-positive: 33.3% (27/81) agreed/strongly agreed that facial-analysis technology would improve pain assessment, while 39.5% (32/81) were neutral. Similarly, 28.4% (23/81) agreed/strongly agreed that it would improve patient care, while 42.0% (34/81) were neutral.

Provider endorsement was higher for targeted applications, especially pain assessment over time. 50.6% (41/81) of providers agreed/strongly agreed that the technology would be useful for monitoring patients over time (Figure 2a), and 45.0% (36/80) agreed/strongly agreed that the technology would help identify patients whose pain is worsening between routine clinical assessments. One clinician added that in the ED, patients “*may act differently when the nurse is in the room vs. not in the room*,” indicating a challenge that could be addressed with continuous monitoring.

**Fig. 2.**
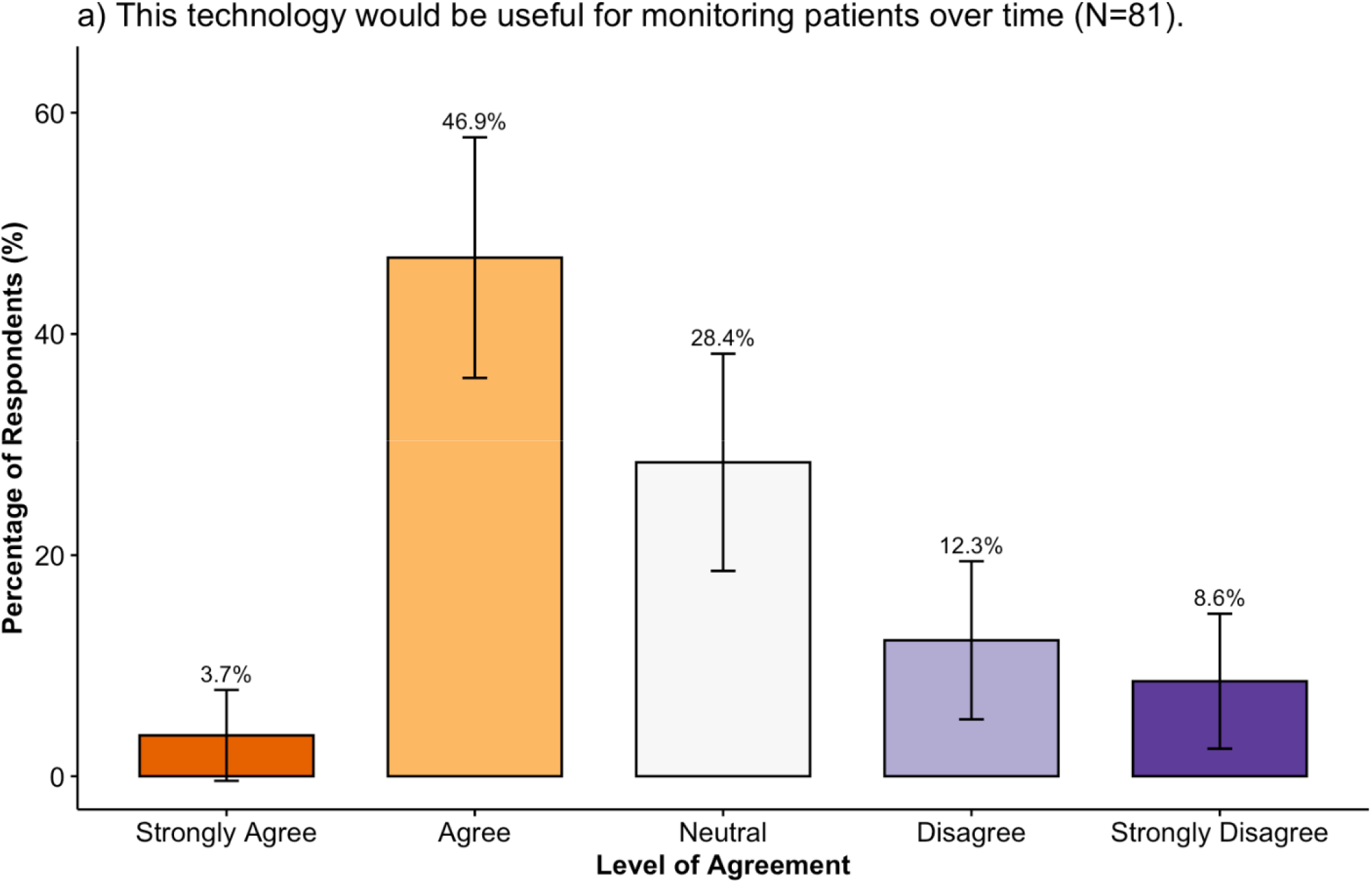
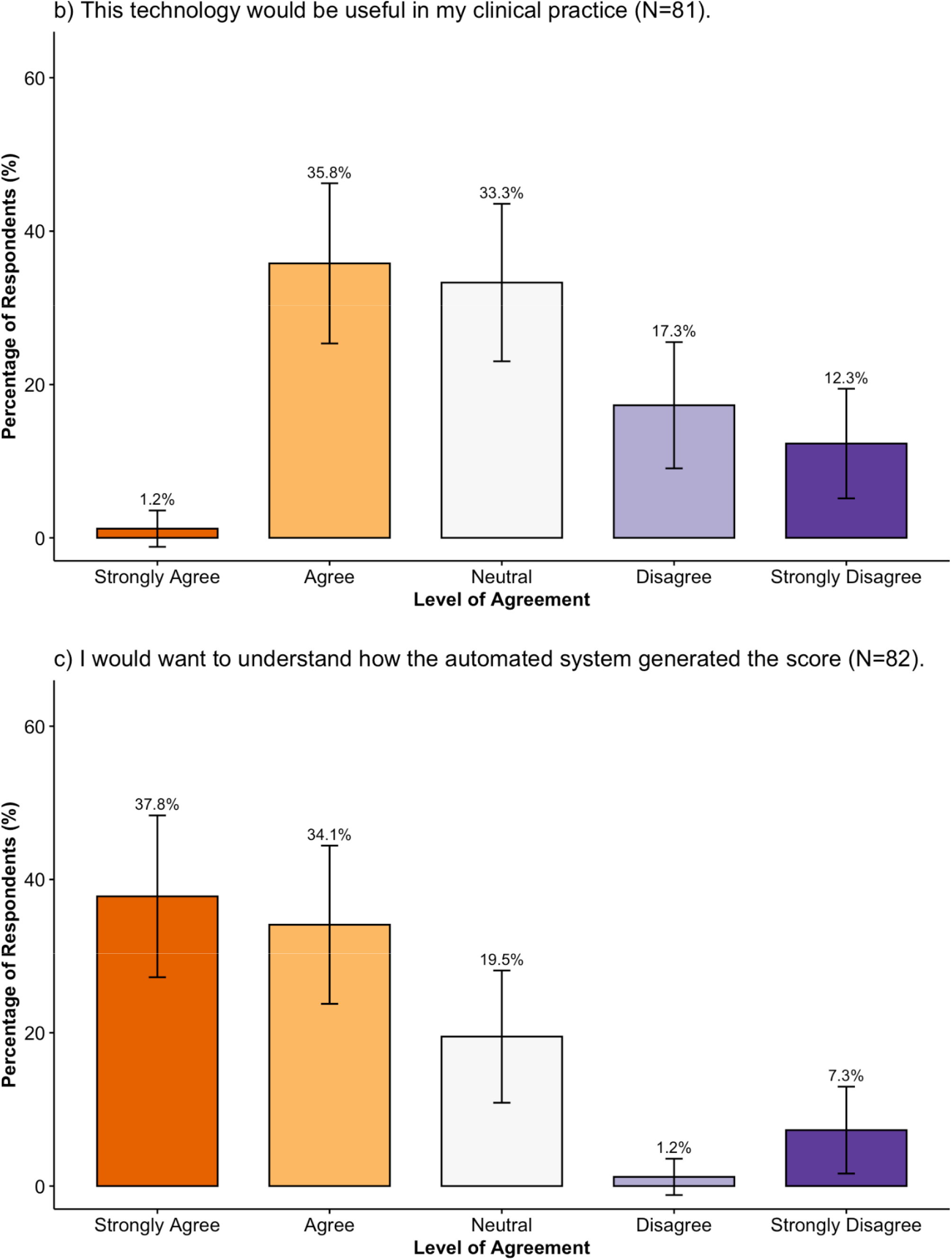
Clinician perspectives on the use of an automated pain detection and scoring system in clinical practice. Figure created in R

Most respondents were neutral (27/81, 33.3%) or agreed/strongly agreed (29/81, 35.8%) that a potential automatic pain recognition and scoring system would be useful in their clinical practic (Figure 2b). A majority of clinicians also agreed/strongly agreed (59/82, 80.0%) that they would want to understand how the system generated the score (Figure 2c). Subgroup analyses demonstrated a significant gender difference in perceived usefulness, with women more likely than men to agree that the technology would be useful in their clinical practice (*p*=0.043). No other significant subgroup differences (clinical role, years experience) were identified.

Clinicians reported the system would be most useful for nonverbal patients (64/74, 86.5%), patients with dementia or cognitive impairments (58/74, 78.4%), and patients with language barriers (46/74, 62.2%; Figure 3). One respondent elaborated, “*For altered or non-verbal patients this has the potential to really aid in bedside pain management*.”

**Fig. 3.**
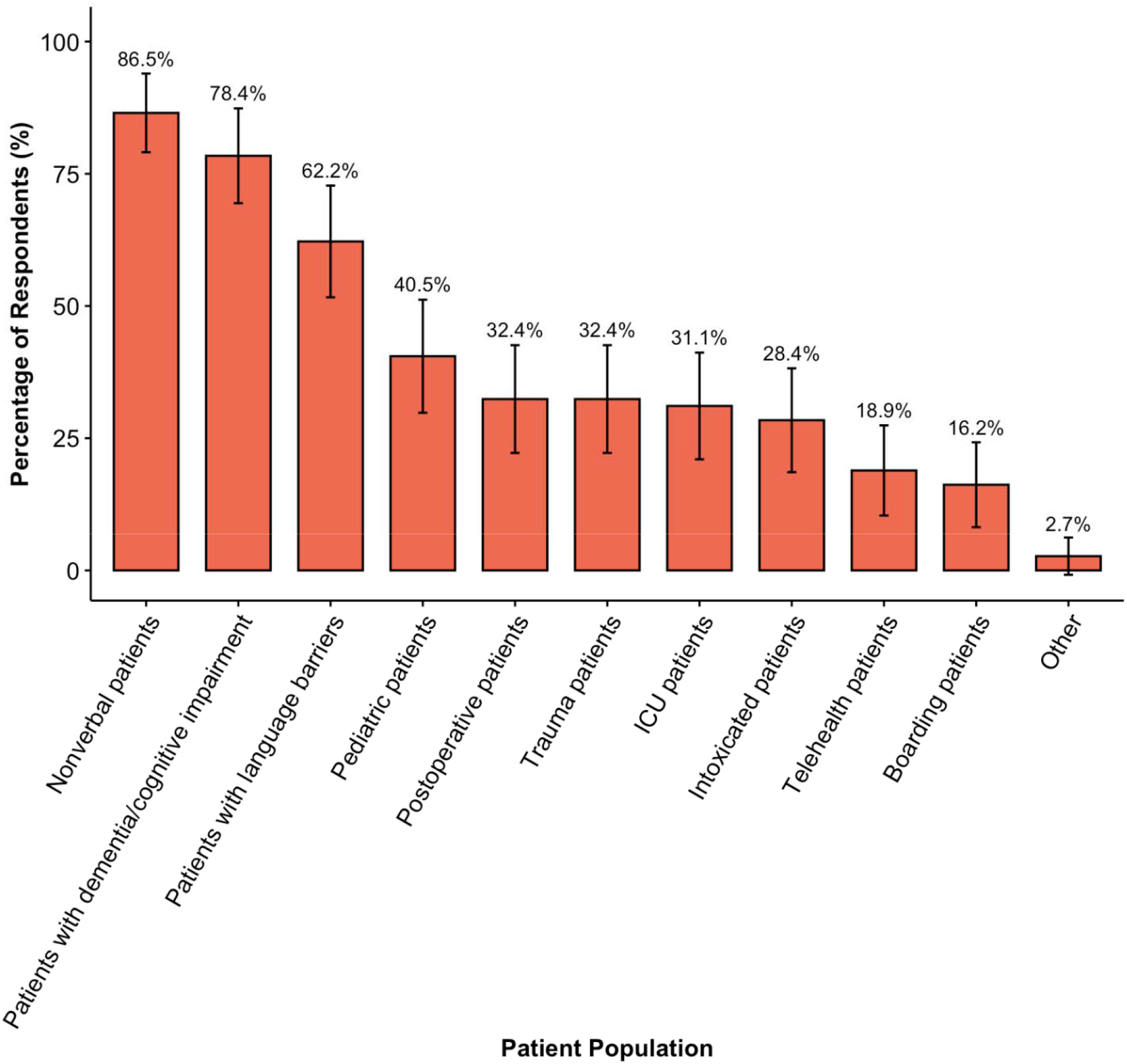
Populations for which an automatic pain detection and scoring system may be useful (N=74). Figure created in R

These perceived benefits mirror challenges clinicians cited in their current pain assessment practices: the greatest challenges were patients unable to communicate verbally (62/87, 71.3%), inconsistent patient self-reporting (59/87, 67.8%), patients with altered mental status (53/87, 60.9%), and time constraints or high patient volume (46/87, 52.9%). In the “Other” free-text field, one respondent elaborated on incongruence between patient-reported pain score and provider assessment:

“*Patient smiling, chatting on phone and then stating 10/10*.”

Another respondent expressed concern regarding patient-reported pain scores, noting that patients “*[know] what number to say to get the highest dose of PRN pain medication*.”

In the free-text responses, clinicians also reported specific patient populations in which the automated system would not be useful:

“*I don’t think this would work for children/babies who cry all the time for various reasons*.” “*In the ICU many of my patients are intubated, I think this will likely be difficult for the system*.”

Provider responses varied in the level of accuracy required to use a potential pain detection and scoring system (Table 2). Almost two-thirds of respondents reported they would be likely/very likely to use the system with at least 90% accuracy (n=53/81, 65.4%). Accuracy requirements differed by clinical role (*p*=0.003): a majority of physicians reported willingness to use the system with at least 80% accuracy, whereas nurses and APPs required 90% or higher accuracy. Subgroup analyses identified gender differences in the accuracy requirement, with women reporting a higher required accuracy than men *p*=0.027). In additional questions about system error, 54.4% (43/79) of clinicians were very or extremely concerned about the system reporting a high pain score when the patient’s pain was low (false-positive), and 56.4% (44/78) were concerned about the system reporting a low pain score when the patient’s pain was high (false-negative). In the free-response text, clinicians indicated a need for rigorous system validation:

**Table 2.**
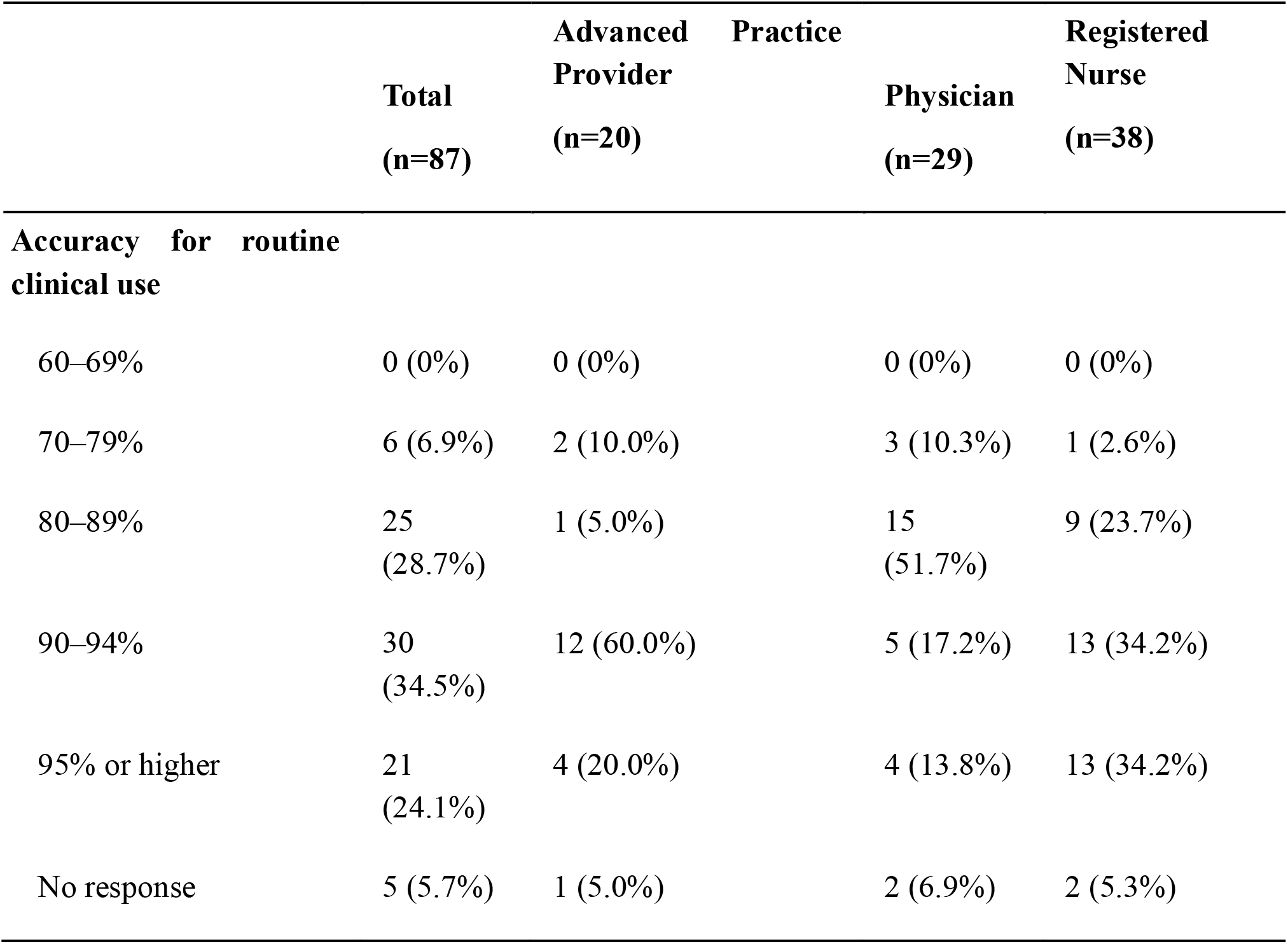
Stated requirements for automatic pain detection and scoring system accuracy by clinical role.

“*Such a system would need to be very well-studied and validated before I would be comfortable using it. I can imagine many potential pitfalls (validity for one diagnosis, but not another, variation in effectiveness based on age, race, gender, variation based on time of day)…*”

We assessed clinician concerns about different aspects of this type of system. Clinicians expressed concerns about excessive alerts or notifications, with 72.2% (57/79) of providers indicating they were very or extremely concerned (Figure 4a). 67.9% (53/78) of respondents also reported being very or extremely concerned about unequal system performance across patient populations (Figure 4b). Clinicians further expressed concern about unequal performance across patient groups in the free-response:

**Fig. 4.**
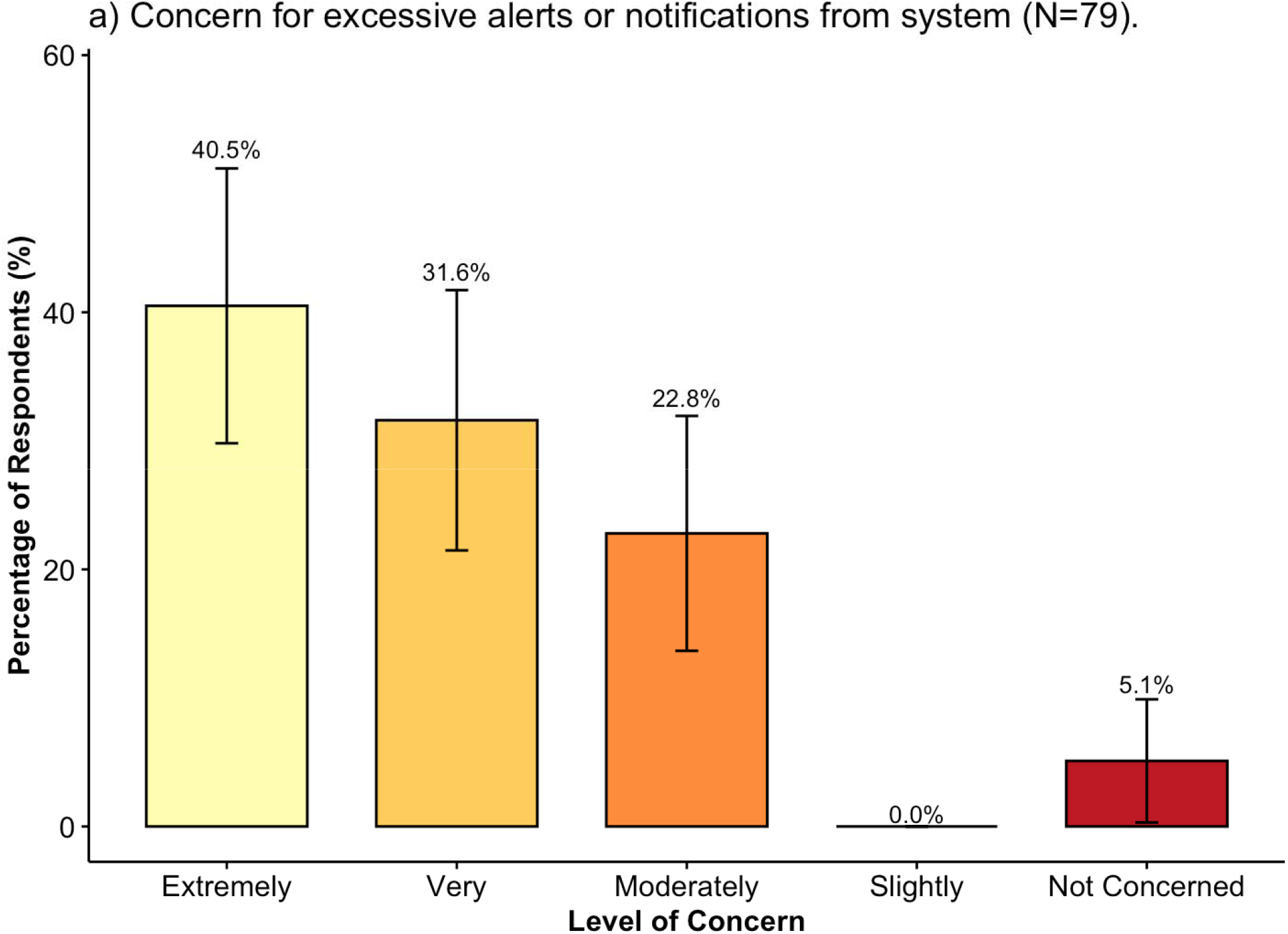
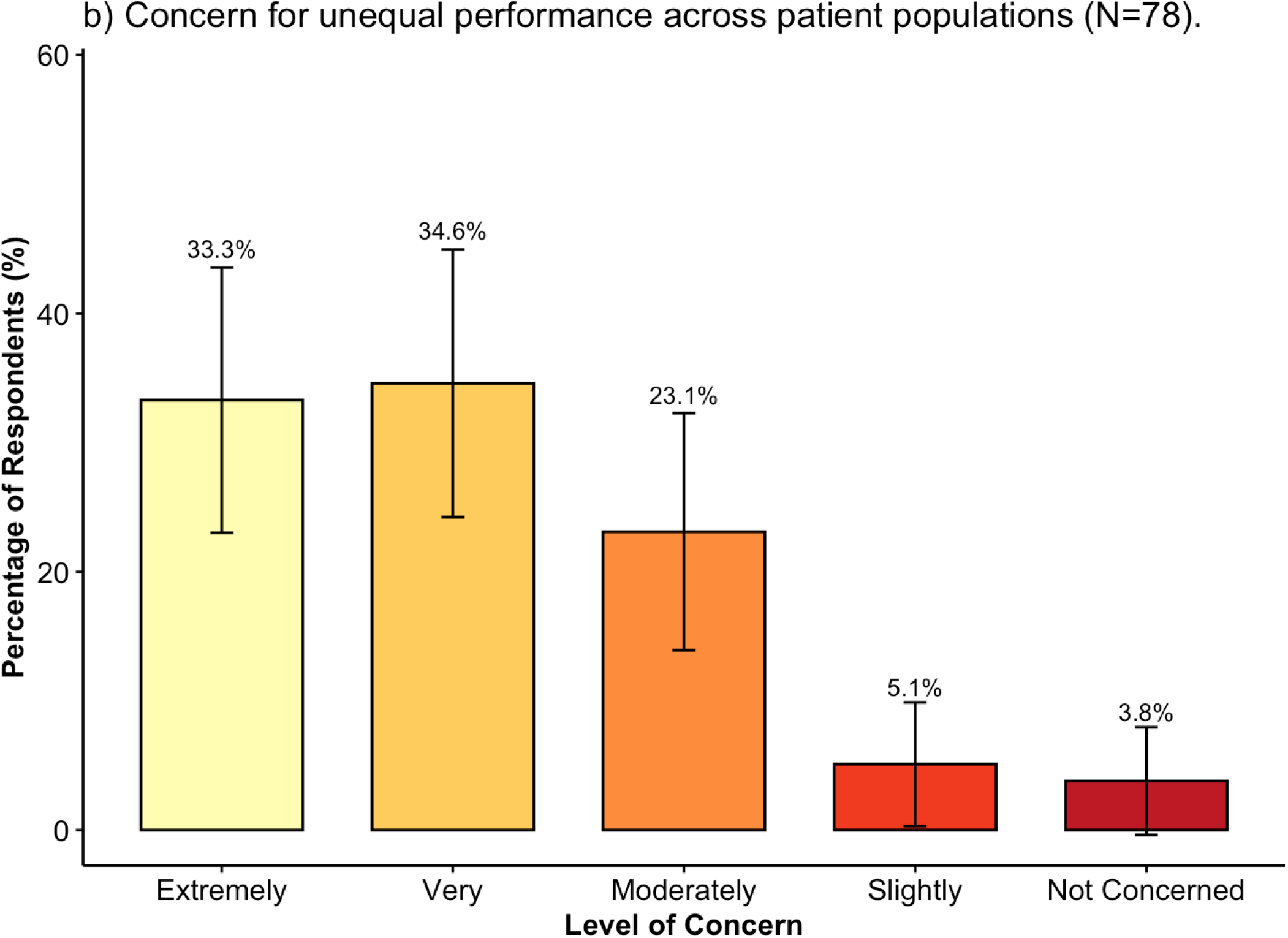
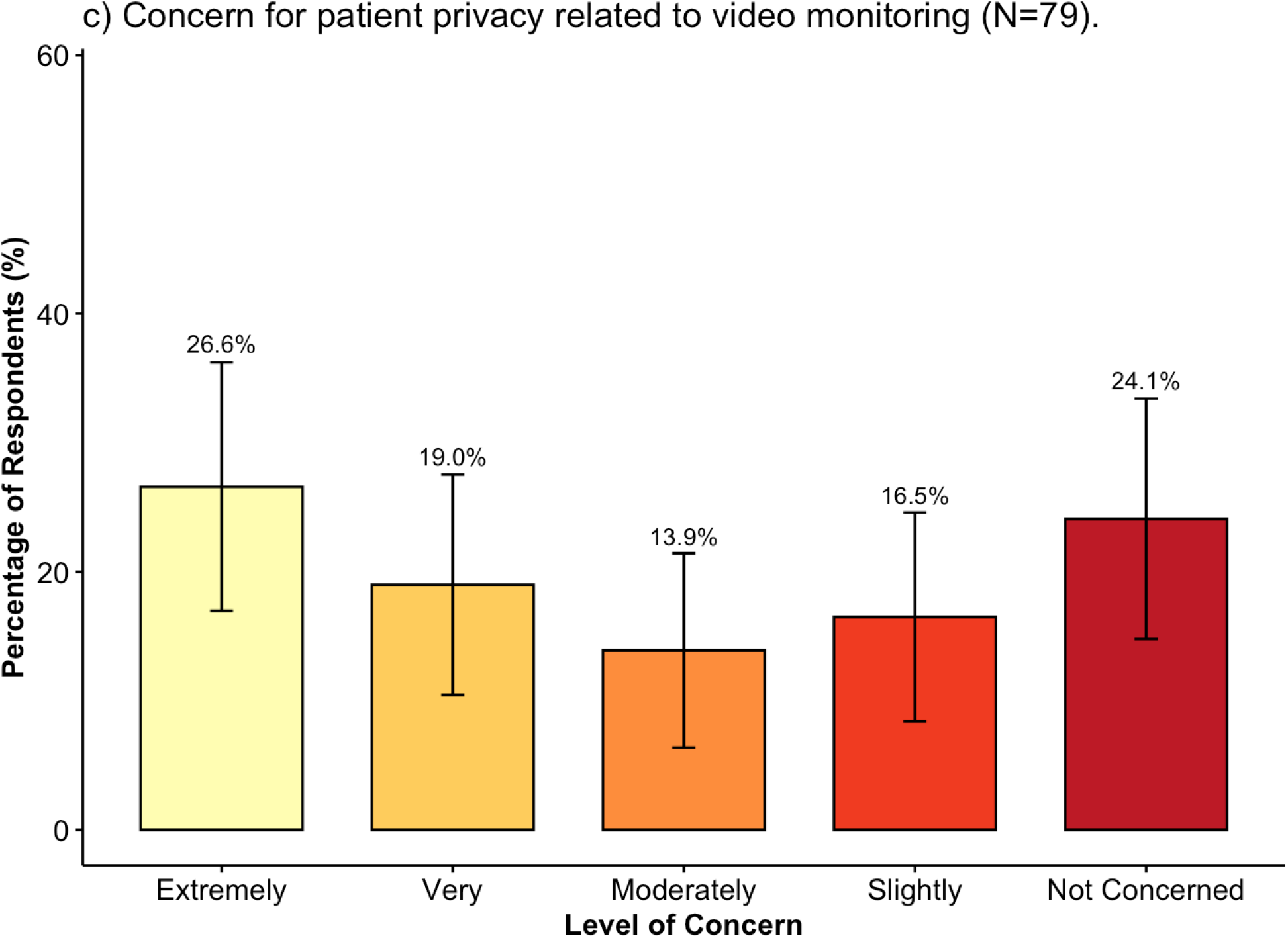
Stated clinician concerns about an automated pain detection and scoring system. Figure created in R

“*I worry about the inherent racial biases with facial recognition*.”

“*Facial expressions are too variable. Will not work correctly for neurodivergent people or people who habitually clench their teeth*.”

Providers had mixed opinions regarding patient privacy concerns related to video monitoring, with 40.5% (32/79) being not concerned or slightly concerned, and 45.6% (36/79) being very or extremely concerned (Figure 4c). The free-response text expanded on these concerns:

“*I’m also weary of data collection and having cameras pointed at patients constantly may make families/patients uncomfortable*.”

Subgroup analyses identified no significant differences in concerns regarding excessive alerts, unequal performance across patient populations, or patient privacy according to clinical role, years in clinical role, or gender.

Additional free-text responses are reported in Supplementary Material 5.

## Discussion

### Survey Participant Characteristics and Data Completeness

This survey study was among the first to explore clinicians’ perspectives on pain assessment practices and the potential role of an automatic pain detection and scoring system centered on facial expressions. Our sample included physicians, advanced practice providers, and nurses across 11 clinical departments, providing a comprehensive insight into different pain assessment contexts and clinical workflows. Participants were also nearly evenly divided by clinical experience (i.e., less than 5 years vs. more than 5 years). These factors expand on existing literature wherein previous studies have typically focused on a single department.^23^ Additionally, to our knowledge, this study is the first to specifically explore clinicians’ attitudes towards automated facial expression analysis as a potential approach to pain detection.

### Current pain assessment practices and challenges

Clinicians relied most heavily on observational information when assessing pain, with vital signs, physical examination findings, and facial expressions emerging as the most frequently reported tools. These findings suggest that routine pain assessment often involves a synthesis of observable patient characteristics rather than a single measure (e.g., the gold standard patient-reported NRS). The prominence of vital signs and physical examination findings supports incorporating physiologic and clinical data into future multimodal pain-monitoring systems. Notably, facial expressions were among the most commonly used observational indicators, supporting the clinical relevance of automated facial-analysis approaches as a component of pain assessment.

In contrast, formal pain scales and structured assessment instruments, like CPOT, were cited much less frequently. This may indicate greater reliance on clinical judgment and gestalt, which likely integrates vital signs, physical examination findings, facial expressions, and overall clinical context.

Among patient-reported measures, numerical rating scales and verbal descriptions of pain were the most commonly utilized tools. However, both ranked below clinician-observed information and depend on a patient’s ability to communicate effectively. Consistent with this limitation, respondents identified nonverbal and cognitively impaired patients as key populations that may benefit from automated pain-monitoring technologies, highlighting the potential role of these systems when traditional self-reported pain assessment is challenging or unavailable.

### Potential benefits of facial-analysis technology for pain assessment

Overall, more clinicians viewed the technology favorably than unfavorably for use in their own clinical practice, although a substantial proportion remained neutral (Figure 2b). This is consistent with a previous study of physicians and nurses in an intensive care unit, in which 50% reported a positive attitude toward automated pain recognition while 30.4% would “maybe” use the technology.^23^ The large neutral group in our study may be particularly important for future implementation efforts, as perceptions may be influenced by demonstrated performance, workflow integration, and clinical experience with the system.

Nearly half of respondents believed that an automated pain assessment system would be most useful for monitoring patients over time (Figure 2a). This suggests that the greatest clinical value may not lie in generating a single objective pain score, but rather in tracking changes within an individual patient over time. Comparing patients with their own baseline may also reduce the risk of algorithmic bias due to differences in pain expression between individuals. Longitudinal monitoring may provide clinicians with additional information about worsening or improving pain between routine clinical assessments. A potential system could be integrated into patient care rooms equipped with ambient intelligence technology,^32,33^ with patient-facing cameras and microphones available for continuous monitoring.

Respondents also expressed a need to understand how the system generated its pain prediction, with more than 70% agreeing or strongly agreeing that they would want insight into the automated process. This finding underscores the importance of transparency and explainability in AI-assisted technology. Clinician acceptance may be enhanced by involving end users throughout development, providing targeted education and hands-on training with a prototype system, and offering case examples to compare system outputs with clinical assessments. Successful implementation will require a collaborative approach, incorporating clinician perspectives throughout system design and deployment.

Perceived benefit varied across patient populations (Figure 3). Clinicians expressed little perceived benefits in using automatic pain detection for telehealth patients and only modest interest for trauma, intensive care unit, and postoperative patients. Additionally, less than half of the respondents identified benefits in pediatric populations. In contrast, interest was substantially higher for patients with language barriers, cognitive impairment such as dementia, and nonverbal patients. Nearly 80% of respondents identified patients with cognitive impairment as a population that may benefit from this technology, while nonverbal patients selected most frequently overall (86.5%).

These findings suggest that clinicians perceive the greatest value of automated pain monitoring in populations for whom traditional self-reported measures are difficult or impossible to obtain. Correspondingly, facial analysis has been widely used for pain detection among patients living with dementia,^34–37^ a growing nonverbal patient population. More broadly, nonverbal patients encompass highly heterogeneous populations beyond individuals living with dementia. This emphasizes the potential role of observational technologies as clinical decision-support tools and the importance of considering the needs of specific patient populations in deployment.

### Concerns about facial-analysis technology for pain assessment

Clinicians expressed high requirements for system accuracy, with most respondents requiring at least 90% accuracy before considering routine use of the system, although physicians tolerated lower performance than nurses (Table 2). This suggests that clinician trust, particularly among nursing staff, will require strong technical performance with explainable inputs and outputs. Concerns about accuracy are well-grounded in the current evidence: a meta-analysis of AI-based pain assessment from facial images reported combined sensitivity and specificity of 98%, but all included studies had at least one domain with high risk of bias, and reported accuracies across the broader literature ranged widely from 0.27 to 0.99.^19^ Concerns about variability in pain expression is also supported by evidence that existing automated systems have also been trained primarily on small cohorts of young and healthy populations, raising questions about generalizability to older adults, patients with dementia who may exhibit emotional blunting, and diverse racial and ethnic groups.^38,39^

Two prominent concerns were related to implementation and system development. First, over 70% of clinicians reported being very or extremely concerned about excessive alerts and notifications (Figure 4a), highlighting the risk of exacerbating alert fatigue with an additional system. Automated pain-monitoring systems should therefore be designed to integrate thoughtfully into clinical workflows and minimize cognitive burden. Passive displays of information may be more acceptable than frequent alerts or electronic health record pop-ups. Overall, implementation must be co-designed with clinicians to increase acceptance and routine use.

Second, over two-thirds of respondents expressed concern regarding unequal performance across patient populations (Figure 4b). This concern is particularly important given well-documented challenges of bias in facial-recognition technologies and pain assessment practices. One strategy to reduce potential bias is to emphasize longitudinal estimates and predictions, focusing on changes within a patient over time rather than cross-sectional comparisons between patients. This strategy is grounded in fundamental statistical estimation theory: assuming that bias in the system is invariant over time (i.e., within a patient encounter), the longitudinal trend estimates will be independent of unobserved confounding and measurement errors.^40,41^ Additionally, prospective validation studies should include diverse and representative patient populations and assess calibration across demographic and clinical groups to ensure equitable performance.

Clinicians’ concerns about patient privacy related to facial recording were mixed, with respondents divided between those reporting little concern and substantial concern. The current study focused on clinician attitudes; thus, further investigation is needed to evaluate patient attitudes toward a video recording based pain detection system. To this end, Katsanis et al.^42^ found that while a majority of US adults were comfortable with facial recognition technology in healthcare, a significant fraction expressed privacy concerns about face-based data, and nearly one-quarter would prefer to opt out of facial imaging components of research.

### Limitations

This was a single-site study conducted at an academic medical center, which may limit generalizability to clinical settings with different patient populations, workflows, or institutional cultures. Most patients in this setting receive care from resident physicians, providing an additional layer of pain monitoring that may not be present at community hospitals. This staffing structure could influence the perceived usefulness of automated pain assessment technology; clinicians in resource-limited settings where fewer providers are available for frequent reassessment^43^ may view such tools differently. Future research should include multi-site studies, including community hospitals and settings with varying patient volumes. Additionally, participants evaluated a potential technology that has not yet to be developed, meaning their perspectives reflect anticipated rather than experiential attitudes.

Overall, missing data were low. However, item nonresponse was higher in later sections of the survey. Although a progress bar was included to set expectations regarding survey length, respondent engagement may have declined as the survey progressed. Clinicians with stronger opinions about AI for pain assessment, whether positive or negative, may have been more motivated to respond, potentially overrepresenting strongly held attitudes toward facial-analysis technology.

## Conclusion

Clinicians expressed neutral-to-positive attitudes toward automated facial expression analysis for pain assessment, with the greatest perceived benefit for monitoring pain over time and for patients who may have difficulties with self-reporting. However, concerns regarding accuracy, generalizability, alert fatigue, and equitable performance highlight important barriers to clinical implementation. These findings support further development and prospective validation of multimodal, interpretable, and longitudinal pain-monitoring systems designed in collaboration with clinicians and evaluated across diverse patient populations.

## Declarations

### Ethics Approval and Consent to Participate

Ethical approval was obtained from the University of Virginia Institutional Review Board for Social and Behavioral Sciences (IRB-SBS) under Protocol Number 8544. Informed consent was obtained from all participants prior to participation.

### Competing interests

The authors declare no competing interests.

Acknowledgements

The authors are grateful to the departmental leadership and administrative staff who were instrumental in disseminating the survey to a wide pool of eligible participants.

### Data Availability

Deidentified data will be made available following reasonable requests made to the corresponding author.

### Contributions

CM - Study concept and design, data collection, data analysis, manuscript writing. CE - Study concept and design, data analysis, manuscript writing. MG – manuscript review. PC - Study concept and design, manuscript editing, supervision.

## Notes

### Competing Interest Statement

The authors have declared no competing interest.

### Author Declarations

IRB of the University of Virginia for Social and Behavioral Sciences gave ethical approval for this work under protocol 8544

