## Supplementary Material for "Clinician Perspectives on Automated Facial Analysis for Pain Assessment: A Cross-Sectional Survey of Physicians, Advanced Practice Providers, and Nurses"

**Supplement**

**Supplement 1**

CHERRIES Checklist

**Supplement 2**

Survey

**Supplement 3**

**Supplementary Table 1: Perceived difficulty to assess and monitor patients over time in clinicians’ current workflow**

|  | **Total**  **(n=87)** |
| --- | --- |
| Extremely easy | 3 (3.4%) |
| Somewhat easy | 31 (35.6%) |
| Neither easy nor difficult | 22 (25.3%) |
| Somewhat difficult | 27 (31.0%) |
| Extremely difficult | 4 (4.6%) |

**Supplementary Table 2: Challenges in accurately assessing patient pain.** Note: participants selected all that apply

|  | **Total**  **(n=87)** |
| --- | --- |
| Frequent handoffs between providers | 25 (4.6%) |
| Inconsistent patient self-reporting | 59 (10.8%) |
| Intoxication or substance use | 29 (5.3%) |
| Lack of continuous monitoring | 19 (3.5%) |
| Language barriers | 42 (7.7%) |
| Limited objective measures of pain | 39 (7.1%) |
| Limited opportunities for reassessment | 37 (6.8%) |
| Altered mental status (e.g., dementia, delirium) | 53 (9.7%) |
| Patient unable to communicate verbally | 62 (11.4%) |
| Psychiatric illness | 25 (4.6%) |
| Time constraints/high patient volume | 46 (8.4%) |
| Other | 4 (0.7%) |

**Supplementary Table 3: Clinician level of agreement with the given statements**

|  | **Strongly Agree** | **Agree** | **Neutral** | **Disagree** | **Strongly Disagree** | **No response** |
| --- | --- | --- | --- | --- | --- | --- |
| I would personally use this technology | 1 (1.1%) | 22 (25.3%) | 32 (36.8%) | 13 (14.9%) | 13 (14.9%) | 6 (6.9%) |
| I would want to understand how the automated system generated the score | 31 (35.6%) | 28 (32.2%) | 16 (18.4%) | 1 (1.1%) | 6 (6.9%) | 5 (5.7%) |
| This technology would be useful for nonverbal or altered patients | 6 (6.9%) | 44 (50.6%) | 19 (21.8%) | 9 (10.3%) | 3 (3.4%) | 6 (6.9%) |
| This technology would be useful for monitoring patients over time. | 3 (3.4%) | 38 (43.7%) | 23 (26.4%) | 10 (11.5%) | 7 (8%) | 6 (6.9%) |
| This technology would be useful in my clinical practice | 1 (1.1%) | 29 (33.3%) | 27 (31%) | 14 (16.1%) | 10 (11.5%) | 6 (6.9%) |
| This technology would help identify patients whose pain is worsening between routine clinical assessments | 3 (3.4%) | 33 (37.9%) | 26 (29.9%) | 11 (12.6%) | 7 (8%) | 7 (8%) |
| This technology would improve pain assessment | 2 (2.3%) | 25 (28.7%) | 32 (36.8%) | 14 (16.1%) | 8 (9.2%) | 6 (6.9%) |
| This technology would improve patient care | 2 (2.3%) | 21 (24.1%) | 34 (39.1%) | 14 (16.1%) | 10 (11.5%) | 6 (6.9%) |

**Supplementary Table 4: Clinician likelihood of using the system at the following levels.**

|  | **Very Likely** | **Likely** | **Neutral** | **Unlikely** | **Very Unlikely** | **No response** |
| --- | --- | --- | --- | --- | --- | --- |
| 60-69% | 0 (0%) | 1 (1.1%) | 11 (12.6%) | 25 (28.7%) | 44 (50.6%) | 6 (6.9%) |
| 70-79% | 0 (0%) | 10 (11.5%) | 14 (16.1%) | 22 (25.3%) | 35 (40.2%) | 6 (6.9%) |
| 80-89% | 7 (8%) | 21 (24.1%) | 21 (24.1%) | 9 (10.3%) | 23 (26.4%) | 6 (6.9%) |
| 90-94% | 18 (20.7%) | 35 (40.2%) | 16 (18.4%) | 3 (3.4%) | 9 (10.3%) | 6 (6.9%) |
| 95% or higher | 41 (47.1%) | 22 (25.3%) | 10 (11.5%) | 5 (5.7%) | 4 (4.6%) | 5 (5.7%) |

**Supplementary Table 5: Clinician likelihood of taking the following actions if the system detected worsening pain.** Note: only physicians and APPs could respond to the “Notify nursing staff” option, and only nurses could respond to the “Notify a physician/APP” option

|  | **Very Likely** | **Likely** | **Neutral** | **Unlikely** | **Very Unlikely** | **No response** |
| --- | --- | --- | --- | --- | --- | --- |
| Consider additional analgesia | 10 (11.5%) | 47 (54%) | 16 (18.4%) | 2 (2.3%) | 2 (2.3%) | 10 (11.5%) |
| Notify a physician/APP | 2 (2.3%) | 21 (24.1%) | 8 (9.2%) | 1 (1.1%) | 1 (1.1%) | 54 (62.1%) |
| Notify nursing staff | 7 (8%) | 24 (27.6%) | 10 (11.5%) | 1 (1.1%) | 1 (1.1%) | 44 (50.6%) |
| Reassess the patient | 21 (24.1%) | 47 (54%) | 6 (6.9%) | 1 (1.1%) | 2 (2.3%) | 10 (11.5%) |

**Supplementary Table 6: Clinician level of concern regarding the following statements**

|  | **Extremely** | **Very** | **Moderately** | **Slightly** | **Not Concerned** | **No response** |
| --- | --- | --- | --- | --- | --- | --- |
| Excessive alerts or notifications from system | 32 (36.8%) | 25 (28.7%) | 18 (20.7%) | 0 (0%) | 4 (4.6%) | 8 (9.2%) |
| Increased workload | 15 (17.2%) | 16 (18.4%) | 14 (16.1%) | 25 (28.7%) | 9 (10.3%) | 8 (9.2%) |
| Legal responsibility related to automated or AI-generated pain score | 24 (27.6%) | 22 (25.3%) | 16 (18.4%) | 9 (10.3%) | 8 (9.2%) | 8 (9.2%) |
| Patient privacy concerns related to video monitoring | 21 (24.1%) | 15 (17.2%) | 11 (12.6%) | 13 (14.9%) | 19 (21.8%) | 8 (9.2%) |
| System gives high pain score when patient is at low pain level | 19 (21.8%) | 24 (27.6%) | 25 (28.7%) | 6 (6.9%) | 5 (5.7%) | 8 (9.2%) |
| System gives low pain score when patient is at high pain level | 20 (23%) | 24 (27.6%) | 24 (27.6%) | 6 (6.9%) | 4 (4.6%) | 9 (10.3%) |
| Unequal performance across patient populations | 26 (29.9%) | 27 (31%) | 18 (20.7%) | 4 (4.6%) | 3 (3.4%) | 9 (10.3%) |

**Supplement 4**

Fisher’s tests for the questions in Supplemental Table 7.

Supplemental Table 7. Questions assessed with Fisher’s tests.

|  | **Clinician level of agreement with the given statements.** |
| --- | --- |
| Q3.2 | I would want to understand how the automated system generated the score. |
| Q3.4 | This technology would be useful for monitoring patients over time. |
| Q3.5 | This technology would be useful in my clinical practice. |
| Q4 | **Clinician likelihood of using the system at the following levels.** |
|  | **Clinician level of concern regarding the following statements.** |
| Q6.1 | Excessive alerts or notifications from system |
| Q6.4 | Patient privacy concerns related to video monitoring. |
| Q6.7 | Unequal performance across patient populations. |

Supplementary Table 8. Fisher’s test p values.

|  | By clinical role | By years in role | By gender |
| --- | --- | --- | --- |
| Q3.2 | 0.536 | 0.515 | 0.832 |
| Q3.4 | 0.373 | 0.315 | 0.415 |
| Q3.5 | 0.054 | 0.465 | 0.043* |
| Q4 | 0.003* | 0.868 | 0.027* |
| Q6.1 | 0.255 | 0.435 | 0.661 |
| Q6.4 | 0.848 | 1.000 | 0.419 |
| Q6.7 | 0.869 | 0.481 | 0.363 |

*Significance at p < 0.05.

**Supplement 5**

**Additional commentary on free-text responses**

Taken as a whole, the responses to the Likert-scale questions were generally neutral-to-positive towards the proposed technology. In contrast, the free-response section reflected more critical and cautionary perspectives. Some clinicians expressed strong opposition to the technology and Artificial Intelligence more broadly, with one stating, “*I hate this idea and personally it would deter me from seeking care at all*,” while another commented, “*I do not trust AI farther than I can throw it*.”

Respondents raised concerns about the potential for patients to manipulate the technology. One respondent noted, “*Some ED populations that abuse the system would be able to take advantage of the tool to inflate their pain answers*.” Concerns regarding overreliance on technology were also expressed, with one clinician stating, “*I worry about the reliance on technology to take over clinical decision making and patient communication and connection.*”

Providers additionally questioned the ability of facial expression to accurately reflect subjective pain experiences:

“*Pain is subjective to the patient and facial recognition is an objective assessment. Just because a patient does look to be in pain or show it on their face, does not mean that they are feeling pain. One person may be calm with neutral expression at 8/10 and another may be grimacing/crying at 8/10*.”

Another respondent stated, “*Patients do not always outwardly express pain, particularly patients with chronic severe pain*.” Respondents also highlighted the difficulty of distinguishing pain from other sources of discomfort: “*With many of our dying patients they have facial expressions of discomfort that may not necessarily be pain but another discomfort including anxiety, agitation, nausea, constipation*.” Several clinicians also highlighted the difficulty of distinguishing pain from other forms of distress in pediatric populations.

Consistent with these concerns, providers emphasized that facial expression represents only one component of pain assessment. One respondent stated that it is “*difficult to assess pain purely based off facial recognition. Many factors are used to fully determine patients’ pain levels.*”

Some clinicians perceived limited added benefit from facial expression analysis beyond their own assessments. One nurse commented, “*I still need to assess my patient before I give them anything for pain, so this technology would change nothing for me…they [doctors] are going to trust my assessment over facial recognition before ordering any medicine*.” Another provider similarly stated, “*I would be hesitant to trust facial recognition over my own visual assessment*.”

Overall, free-text responses reflected several areas of concern regarding the use of facial expression analysis for pain assessment, particularly regarding trust, the potential for overreliance or manipulation, and the variability of pain expression.
